# COVID-19: Modelling Local Transmission and Morbidity effects to provide an estimate of overall Relative Healthcare Resource Impact by General Practice Granularity

**DOI:** 10.1101/2020.03.20.20039024

**Authors:** Michael Stedman, Mark Lunt, Mark Davies, Martin Gibson, Adrian Heald

**Affiliations:** Res Consortium, Andover, Hampshire; Department of Diabetes and Endocrinology, Salford; The Faculty of Biology, Medicine and Health and Manchester Academic Health Sciences Centre, University of Manchester

**Author notes:** Corresponding author: Dr Mark Davies, Res Consortium, Fosse House, East Anton Court, Icknield Way, Andover, Hants, SP10 5RG.

**Keywords:** COVID-19, general practice, resources, workforce

## Abstract

**Introduction:** Severe Acute Respiratory Syndrome Coronavirus-2 (SARS-CoV-2) is the name given to the 2019 novel coronavirus. COVID-19 is the name given to the disease associated with the virus. SARS-CoV-2 is a new strain of coronavirus that has not been previously identified in humans.

**Methods:** Two key factors were analysed which when multiplied together would give an estimate of relative demand on healthcare utilisation. These factors were case incidence and case morbidity.

GP Practice data was used as this provided the most geographically granular source of published public population data. To analyse case incidence, the latest values for indicators that could be associated with infection transmission rates were collected from the Office of National Statistics (ONS) and Quality Outcome Framework (QOF) sources. These included population density, % age >16 at fulltime work/education, % age over 60, % BME ethnicity, social deprivation as IMD 2019, Location as latitude/longitude, and patient engagement as % self-confident in their own long term condition management.

Average case morbidity was calculated by applying the international mortality Odds Ratio to the local population relevant age and disease prevalences and then summing and dividing by the equivalent national figure. To provide a comparative measure of overall healthcare resource impact, individual GP practice impact scores were compared against the median practice.

**Results:** The case incidence regression is a dynamic situation with the significance of specific factors moderating over time as the balance between external infection, community transmission and impact of mitigation measures feeds through to the number of cases. It showed that currently Urban, % Working and age >60 were the strongest determinants of case incidence.

The local population comorbidity remains unchanged. The range of relative HC impact was wide with 80% of practices falling between 20%-250% of the national median.

Once practice population numbers were included it showed that the top 33% of GP practices supporting 45% of the patient population would require 68% of COVID-19 healthcare resources. The model provides useful information about the relative impact of Covid-19 on healthcare workload at GP practice granularity in all parts of England.

**Conclusion:** Covid-19 is impacting on the utilisation of health and social care resources across the country. This model provides a method for predicting relative local levels of disease burden based on defined criteria and thereby providing a method for targeting limited (and perhaps soon to be scarce) care resources to optimise national, regional and local responses to the COVID-19 outbreak..

## Introduction

This coronavirus, first detected in China in 2019, is genetically closely related to the SARS-CoV-1 virus. SARS emerged at the end of 2002 in China, and it caused more than 8000 cases in 33 countries over eight months. Around one in ten of the people who developed SARS died^1^.

At the date of the submission of this article, the COVID-19 outbreak has caused a total of around 142 539 confirmed (9769 new) cases reported globally with around 1200 in the UK alone. Of these proven cases, around 5393 deaths have been reported due to the virus (around 3.8% mortality rate).^2^ Unlike influenza, there is no vaccine and no specific treatment for the disease.

The implications for countries are that there is likely to be unprecedented demands on all aspects of health and social care resources. In the UK emergency planning is well underway although a key challenge is the capacity of secondary care to manage admissions of unwell patients and of primary care (GP practices) to manage those who not requiring hospital admission.^3^ During an outbreak, decision-makers face surges in resource demand which require resource prioritisation and re-allocation^4^. Predicting areas of greater need is therefore critical to optimise the value of limited resources and funding in the response to the COVID-19 pandemic.

Response to infectious diseases are carried out in different phases, the Government has recently under the advice of Public Health England moved the disease from containment strategy to delay strategy where slowing the onward transmission rate is the priority. Later stages might require other strategy changes.

This paper describes the development of a resource allocation prediction model for COVID-19 based on a regression analysis of published case rates in England across upper-tier local authority areas (UTLA) against key local population metrics aggregated up from local GP practices.

It is worth noting that these published case numbers are only those that have tested positive either from existing patient contacts or sufficiently symptomatic to be triaged to healthcare services. Around 3% of those tested show positive results, while it is estimated that unreported cases could be more than 20 times higher than reported, however the latter remain the only marker for local levels of the condition. The situation is also changing on a real-time basis as the case number grow and moves from initial infection to community transmission, and the mitigation measures start to work through.

## Methods

Two key factors were analysed which when multiplied together would give an estimate of relative demand on healthcare utilisation. These factors were *case incidence* and case *morbidity*. GP Practice data was used as this provided the most geographically granular source of published public population data. To analyse case incidence, the latest values for indicators that could be associated with infection transmission rates were collected from the Office of National Statistics (ONS) and Quality Outcome Framework (QOF) sources. These local factors included population density, % age >16 at fulltime work/education, % age over 60, % BME ethnicity, social deprivation as IMD 2019, Location as latitude and longitude, and patient engagement level as % self-confident in their own long-term condition management.

A dataset was then created by aggregating GP practice data within each UTLA. A stepwise regression analysis was then performed linking these metrics against the latest number of identified cases of COVID-19 in each UTLA area as published by Public Health England^5^ divided by the total population. The factors are inter-related and the total number of factors was kept small while maximising the model variation capture

Applying the regression coefficients for the chosen indicators to actual practice values gave relative incidence values for expected local cases/population, this was divided by the same calculation using national median practice values to show the practice relative incidence rate as % of the national median.

Relative case morbidity was calculated by applying the published Odds Ratio (OR)^6^ analysis for international COVID data on mortality by the local GP practice population percentage prevalence for patient age groups, along with comorbidities including diabetes, coronary heart disease, chronic obstructive pulmonary disease (COPD) and cancer, and then divide by the same calculated value using national average values to give a relative practice relative case morbidity for Covid-19 infection.

Multiplying the practice relative incidence rate by practice relative case morbidity gave the relative practice health care resource impact /patient to the national median. Multiplying this by practice population size divided by the national median practice size gave the total health care resource prediction relative to the median practice.

## Results

The situation is very dynamic and fluid with the number of cases being published daily, the first publication on 8^th^ March 2020 gave 224 cases in 74 UTLAs. The chosen factors and their significance within the statistical model are also changing with time.

The current regression model data was updated on the date of submission of this paper (16.03.2020) from 149 UTLAs of which 136 had recorded one or more out of a total of 1,421 cases in England. The analysis of COVID-19 incidence of infection showed significant positive relation to three key factors: *population density, % of the >16 population in full-time at work/education*, and *% of the population over 60* (r^2^=0.45 and all p-values<0.05).

**Table 1:** Regression results Linking Reported 1,421 COVID-19 Cases on 16-3-2020/,000 population to 3 selected factors in 149 UTLAs with 60 million population.

| Parameter | Mean Value | Median Value | Estimate Coeff | p-value | Standardised beta |
| --- | --- | --- | --- | --- | --- |
| UTLA Population | 400,053 | 309,923 |  |  |  |
| Actual Cases/,000 population |  | .024 | .014 |  |  |
| Constant |  |  | -0.10351 |  | 0 |
| Urban/Rural (Population/ sq km) | 3,131 | 1,925 | 7.47E-06 | <0.0001 | 0.75 |
| >16 in FT Work/Edu as % Pop | 51.9% | 51.0% | 0.137 | 0.036 | 0.18 |
| Age %>60 | 17.8% | 18.2% | 0.155 | 0.013 | 0.26 |

The relative accuracy of the model can be seen in Figure 1 where the model and actual outcomes of cases/population for each UTLA are divided into 3 terciles. 50% of model’s values fall in the same tercile as the actual, 41% in an adjacent tercile, 9% with 2 tercile difference. (If a model contained no link these values would be 33%, 45%, 22%, respectively).

**Fig 1.**
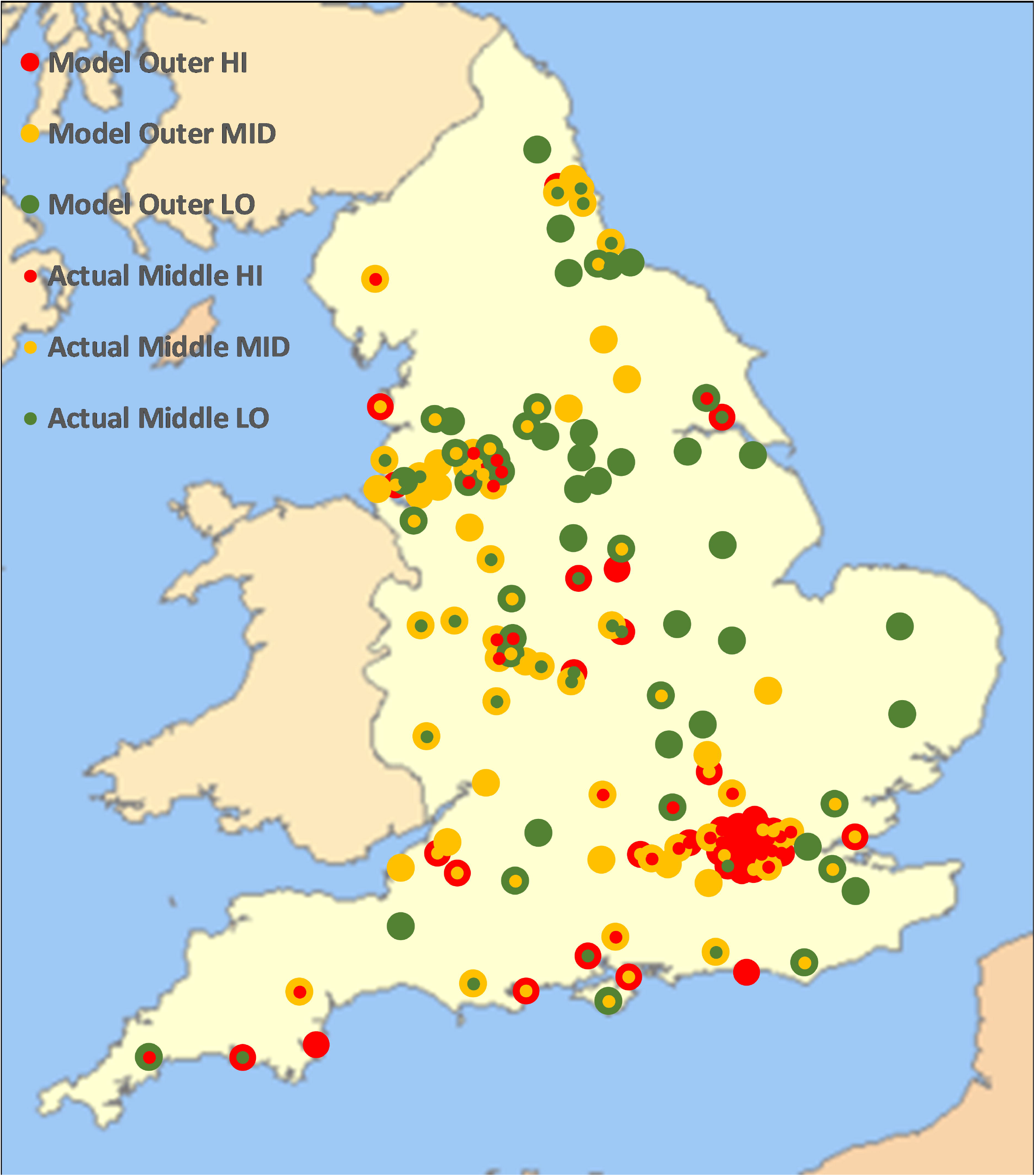
Ranking of Model and Actual Cases/Population taken on 16th March 2020 in each UTLA plotted geographically divided into terciles and shown with the model colour on the outer and the actual colour on the inner

**Figure 2:**
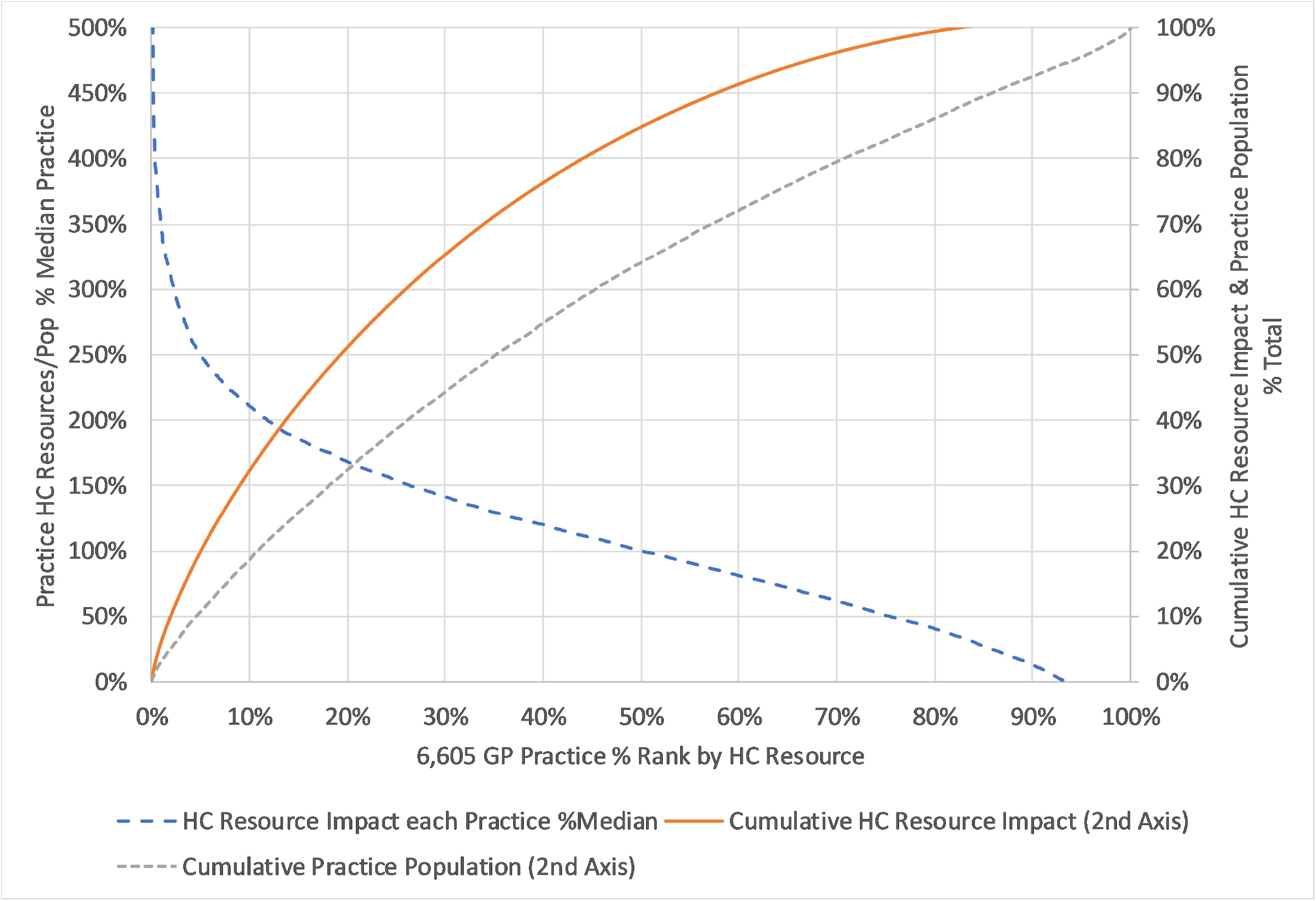
GP Practice Health Resource Impact Model combining expected local case frequency, case comorbidity and practice size.

The application of the extrapolation using regression coefficients and OR combined with each practice’s actual population characteristics show a wide variation in expected cases and overall local morbidity. Combining these two gives a measure of overall expected COVID healthcare resource impact, and this divided by the median GP practice value to give a relative. In figure 1 below, the median practice is shown as 100% of relative healthcare impact, with the highest 10% of GP practices >210% and lowest 10% having <15% of the median. It was multiplied by the relative practice population number and summed cumulatively to show the top 33% of GP practices supporting 45% of the patient population would require 68% of COVID-19 healthcare resources. Specific practices at risk can be identified by their position on these curves.

The authors note that two additional datasets, if and when available, could enhance the predictive capability of the model: local total number of tests and split of tests by numbers and result by known associate or community to give estimates for the level of non-detected, self-managed, populations.

The model both in factor inclusion and resulting values will have to be adjusted on an ongoing basis as the epidemic goes through the recognised infection phases

## Conclusion

The COVID19 outbreak is rapidly impacting on the utilisation of health and social care resources across the country. Linking local case data to local population characteristics provides a method to estimate the difference in expected levels of disease between different populations at the required levels

The factors identified as relevant in the current regression model align with the current strategy of increasing work from home and closing education and reducing social contact especially among older people which should reduce transmission rates. As the situation develops, the transmission drivers will modulate and as further data is available the model can be reviewed, evolved, updated and reissued.

This model provides a method for predicting relative local levels of disease burden based on defined criteria and thereby providing a method for targeting limited (and perhaps soon to be scarce) care resources to optimise national, regional and local responses to the COVID-19 outbreak. We hope that our model will aid precious resource allocation at this very challenging time.

## Data Availability

All the data used is publically available and published on the internet

